# Results from Canton Grisons of Switzerland Suggest Repetitive Testing Reduces SARS-CoV-2 Incidence (February-March 2021)

**DOI:** 10.1101/2021.07.13.21259739

**Authors:** Hossein Gorji, Ivan Lunati, Fabian Rudolf, Beatriz Vidondo, Wolf-Dietrich Hardt, Patrick Jenny, Doortje Engel, Jörg Schneider, Marina Jamnicki, Rudolf Leuthold, Lorenz Risch, Martin Risch, Martin Bühler, Adrian Sommer, Alexa Caduff

**Affiliations:** Laboratory of Multiscale Studies in Building Physics, Empa, Dübendorf, Switzerland; D-BSSE ETH Zürich & Swiss Institute of Bioinformatics, Basel, Switzerland and Federal Office of Public Health FOPH, 3097 Bern, Switzerland; Division of Communicable Diseases, Swiss Federal Office of Public Health, Bern, Switzerland and Institute of Social and Preventive Medicine, University of Bern, Bern, Switzerland; Institute of Microbiology, D-BIOL, ETH Zürich, Switzerland; Department of Mechanical and Process Engineering, ETH Zürich, Switzerland; Department of Justice, Security and Health, Canton Grisons, Switzerland; Clinical Microbiology, Labormedizinisches Zentrum Dr. Risch, 9470 Buchs SG, Switzerland; Faculty of Medical Sciences, Private University of the Principality of Liechtenstein, 9495 Triesen, Liechtenstein

## Abstract

In February 2021, in response to emergence of more transmissible SARS-CoV-2 virus variants, the Canton Grisons launched a unique RNA mass testing program targeting the labour force in local businesses. Employees were offered weekly tests free of charge and on a voluntary basis. If tested positive, they were required to self-isolate for ten days and their contacts were subjected to daily testing at work. Thereby, the quarantine of contact persons could be waved. Here, we evaluate the effects of the testing program on the tested cohorts. We examined 121’364 test results from 27’514 participants during February-March 2021. By distinguishing different cohorts of employees, we observe a noticeable decrease in the test positivity rate and a statistically significant reduction in the associated incidence rate over the considered period. The reduction in the latter ranges between 18%-50%. The variability is partly explained by different exposures to exogenous infection sources (e.g., contacts with visiting tourists or cross-border commuters). Our analysis provides the first empirical evidence that applying repetitive mass testing to a real population over an extended period of time can prevent spread of COVID-19 pandemic. However, to overcome logistic, uptake, and adherence challenges it is important that the program is carefully designed and that disease incursion from the population outside of the program is considered and controlled.

## Introduction

### State of knowledge in mass testing

Repetitive testing of people without noticeable symptoms has been proposed as a public health measure in response to COVID-19 pandemic. The concept relies on reducing the effective infectiousness period by isolating the positively tested individuals when they are presymptomatic or asymptomatic. Theoretical studies have demonstrated that repetitive mass testing helps contain the virus spread (*1–4*). They suggest that this strategy can contribute to control the local epidemics and might even provide an alternative to extreme interventions with higher social, psychological, or economic costs, such as blanket lockdowns. However, due to logistic challenges and high costs, skepticism still remains against repetitive testing as its benefits haven’t been proven broadly yet (*5, 6*).

In the current pandemic, several types of SARS-CoV-2 tests with acceptable sensitivity for symptomatic cases have proven successful as diagnostic tools (*7*). For repetitive population screening, however, specificity, test-to-notice time, and sensitivity to asymptomatic virus carriers are crucial, as they may hinder the effectivity of mass testing programs, if not chosen properly (*5*). In addition, the behavior of the population may jeopardize the positive effects of testing if, for instance, participants do not correctly comply with instructions (i.e. not consistently isolating if tested positive) (*5, 6*). Finally, the success of mass testing depends on the possibility to cover a sufficiently large fraction of the population and their contacts. So far, no empirical study has evaluated the benefits of mass testing when applied to a real population over an extended period of time. There is still lack of field studies of sufficient coverage demonstrating that logistic, uptake and adherence challenges can be overcome.

To date, single, not weekly repetitive, population-based mass testing has been carried out and documented in countries such as the UK, China, South Korea, Austria, Luxembourg, and Slovakia, that have mostly used rapid antigen tests. In Slovakia, a few rounds of population-wide mass testing were estimated to yield a 70% decline of infection prevalence (*8, 9*). Pilot mass testing based on Lateral Flow Devices (LFDs) has been conducted in Liverpool, where lower uptake compared to Slovakia’s was found (only 25% of the population was tested in a four-week period) (*10, 11*). Here we report new evidence from a program in the Canton Grisons (Switzerland) suggesting that repetitive virus RNA mass testing is effective in reducing SARS-CoV-2 incidence. Our observational study gives further insight into effectivity of mass testing in presence of a significant number of exogenous contacts resulting from tourists and commuting labor force.

### Test concept in Canton Grisons

The Canton of the Grisons, the largest in surface and easternmost Canton of Switzerland, shares borders with Italy, Austria and Liechtenstein, and has approximately 200’000 inhabitants. A large share of its economy is dedicated to tourism, e.g. in the past winter season, the Canton Grisons received around 200’000 tourists with 20% from abroad. Due to a high proportion of daily to weekly border crossing employees from the neighbouring countries, border disease control measures are less restrictive than in other parts of the world (in particular after the relaxation of the initial lockdown during March-May 2020). The high incidence and death rates of COVID-19 both in Switzerland and in the neighbouring countries during October-November 2020 and the spread of the new variants (especially B 1.1.7. in Switzerland before Christmas and B.1.351 in Tirol in January 2021), urged the Canton Grisons to launch a population-based mass testing campaign in order to intensify their mitigation strategy while maintaining cross-border socio-economical relationships.

Based on the experience acquired from a few rounds of pilot mass testing conducted in selected municipalities, a repetitive testing strategy for employees was developed for the entire Canton. In the first week of February 2021, 174 companies were recruited, with approximately 100 businesses being added every week since then. The decision to focus on the working force was taken to maximize the benefits by targeting individuals with high mobility and large network of contacts (such as hotel employees), and to enable business continuity by preventing outbreaks in professional networks.

The mass testing campaign relies on voluntary repetitive testing of employees, who are mostly tested once per week (twice per week in a few cases). If the test turns positive, employees are asked to self-isolate, while their work contacts are identified and offered daily testing for ten consecutive days. As long as they remain negative, they are allowed to continue to work on company’s premises with few restrictions (e.g. wearing mask and minimizing contacts when possible): they are asked to self-isolate only if their own test result is positive. This “test-and-release” protocol minimizes the number of people in quarantine and thereby limits possible burdens on the participating businesses. In addition, external contacts (e.g., family and friends) of the infected employee are informed to take precautionary measures and test according to the associated contact tracing program. Even though they are not included in the mass testing program, their risk of infection will be reduced by having a part of their contact network (the employees enrolled in the mass testing program) regularly tested and a beneficial secondary effect is expected for the general canton’s population.

The collected saliva samples are analyzed by the Reverse Transcription Polymerase Chain Reaction (RT-PCR) methods. The use of saliva samples ensures convenience of participants during the testing process. By quantifying the operative characteristics of the tests, a high specificity of 0.999873 (corresponds to 1 false positive in 7’900 tests) and sensitivity close to nasopharyngeal swab RT-PCR were found (*12*). Test processing is expedited by mixing the pools of 5 samples (5 to 1) by the automated platform in the laboratory. The results are communicated within 24 hours, resulting in a test-to-notice time of around 1 day (see *Supplementary materials, Study design and data acquisition*).

In total, 1’022 businesses, operating on 1’358 sites, joined the program from the beginning of February until the end of March, 2021. A total of 121’364 tests were conducted among 27’514 employees without symptoms. Positive test results were found in 215 cases (0.78%) (see *Supplementary materials, Data pre-processing*).

### Factors affecting evaluation and how to quantify

Quantifying the effect of such a repetitive mass testing program in businesses in a real-time setting is not straightforward.

On the one hand, estimates of the population-based indicators to quantify the effect size on the epidemic dynamics, such as the reproductive number, are confounded by the number of tests performed which was not constant over the considered period (*13, 14*).

On the other hand, the epidemiological situation changed over time. The proportion of the B.1.1.7 variant increased as a result of its higher transmissibility, whereas the population aged above 80 was being immunized during the period of our analysis, reducing the size of the susceptible population.

Finally, different business sectors (e.g. tourism, banking, construction, with different number of clients and cross-border employees) are present at varying proportions over time. This might significantly influence the effectiveness of the program, since companies with a higher number of customer contacts or cross-border employees are prone to a higher disease incursion.

For these reasons, instead of focusing on population-based epidemiological quantities in the Canton, here we limit ourselves to a finer-grain analysis: we evaluate the impact of testing on the evolution of the epidemics among participants. Therefore, we consider two groups of employees: the first group consists of the newly enrolled ones, the second group of the ones who have been enrolled in the program for a longer time and have therefore been tested repeatedly. The testing results of the newly enrolled participants are used as a proxy for the spread of infection among the population outside of the program (control group without testing). To account for the influence of different epidemiological conditions at the time of enrollment, which change over time, we divided the repeatedly tested employees into different cohorts according to the week during which they joined the program (i.e., we define week 1, week 2, and week 3 cohorts) (see *Supplemenraty materials, Cohort definition*). Here, we analyze the first two months of this program until the Easter break period, for which the data is already consolidated.

## Results

### Test positivity rate and incidence rate ratio

To infer the epidemiological changes among tested employees, we compute two complementary quantities: test positivity rate (TPR) and incidence rate ratio. The seven-day average TPR is calculated as the proportion of new positive cases to the total number of RT-PCR tests per seven days. Figure 1 shows the Loess fit of the seven-day average TPR for the newly enrolled population (black line) along with the same quantity for repeatedly tested cohorts (blue, green and pink lines), (see *Supplementary materials, Overview of statistical methods*). We observe an increase in TPR among newly enrolled employees during the month of February, which is consistent with the increase in frequency of outbreaks in schools registered in this region, as well as with a concurrent expansion of the pandemic in several Swiss regions. The increase of TPR among the newly enrolled population results in quite different upstream epidemiological conditions for the different tested cohorts. Especially, the initial TPR value of the week 3 cohort is twice as high as the one corresponding to the week 1 cohort. Even though TPR was lower during enrollment of the week 1 cohort, this cohort shows a remarkably smaller and more uncertain reduction in TPR than the other two cohorts joining in later weeks.

**Figure 1:**
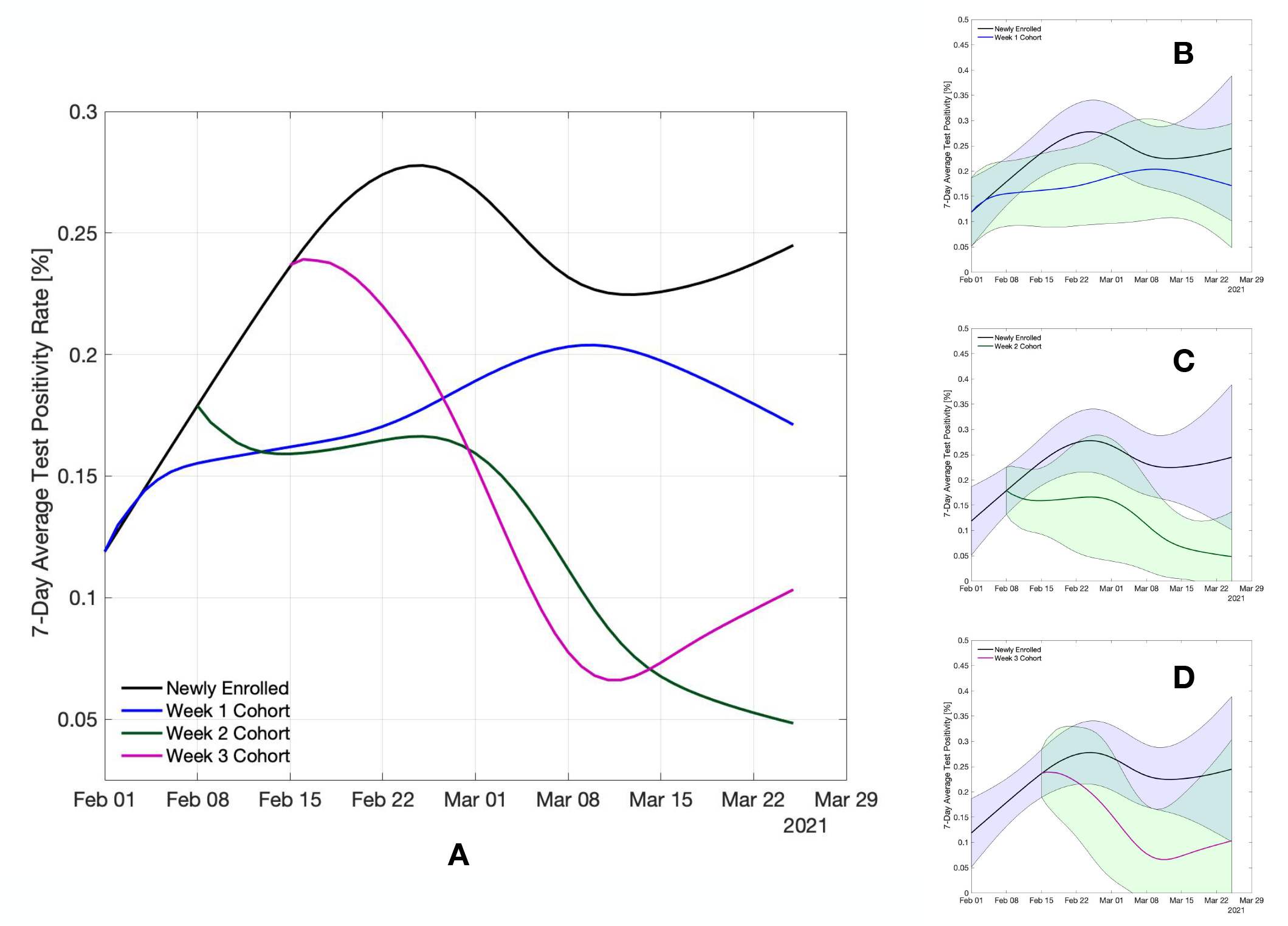
Evolution of TPR (%) for the newly enrolled population as well as other three cohorts considered. Panel A: Loess fitted 7-day average TPR. Panels B, C and D: Loess fitted 7-day average TPR overlaid by 95% CI computed via bootstrapping of the data set (see Fig 4 in *Supplementary materials* for non-smoothed 7-day TPR).

The reduction of TPR over the period in which the cohort is repeatedly tested varies from 73% (over 7 weeks) to 56% (over 6 weeks), for week 2 and week 3 cohorts, respectively. The week 1 cohort shows a 44% increase over the entire eight-week period. Notice that during the same eight-week period, TPR of the newly enrolled population almost doubles (reaching a peak of 0.28% in the fourth week of the program). In general, all three repeatedly tested cohorts of employees exhibit evident reductions in TPR when compared to the newly enrolled population. Yet given the relatively important uncertainty resulting from our small sample size, the critical question is if the reductions in TPR translate into a statistically significant drop in the incidence rate of the tested cohorts.

As an integral measure of the reduction in the number of infections over the entire period, we calculate the incidence rate associated with the seven-day average TPR by normalizing the number of new positive cases by the person-time tested (see *Supplementary materials, Overview of statistical methods*). The ratio between the incidence rate of each cohort with respect to the newly enrolled population is provided in Table 1. The p-values reported in the table, estimated by mid-p exact method, correspond to the test that the incidence rate in the considered cohort has a lower value than the one in the newly enrolled population, hence testing is effective in reducing the number of infections. The estimated reduction in the incidence rate is 18% (95% CI: 0-47%) for the week 1 cohort, 47% (95% CI: 5-71%) for the week 2 cohort and 50% (95% CI: 4-76%) for the week 3 cohort. For the week 2 and week 3 cohorts, we find a statistically significant reduction of the incidence rate achieved by repetitive testing (p-values 0.03 and 0.04, respectively).

**Table 1:** Ratio between the incidence rate of each repeatedly tested cohort with respect to the newly enrolled population

| Starting week of cohort | Estimate | p-value | 95% CI |
| --- | --- | --- | --- |
| week 1 | 0.82 | 0.36 | (0.53,1.25) |
| week 2 | 0.53 | 0.03 | (0.28, 0.95) |
| week 3 | 0.50 | 0.04 | (0.24, 0.96) |

The milder reduction in TPR and incidence rate of the week 1 cohort can partly be explained by a higher exposure to exogenous disease incursion due to the higher representation of the tourism sector and cross-border employees in week 1 cohort in comparison to week 2 and week 3 cohorts (for more details see also *Supplementary materials, Effect of tourism and cross-border commuters*).

### Tourist season and exogenous contacts

Tourism industry is one of the major economical resources of the Canton Grisons. During the past winter season (December 2020 to March 2021), around 200’000 tourists resulted in an additional exogenous infection source in the Canton. Around a quarter (26%) of all tests were carried out among people employed in the tourism sector. Remarkably, this business sector accounted for almost half (48%) of all positive tests, which results in a frequency of positive cases that is close to two times larger than in other business sectors.

Figure 2 indicates that approximately 45% of the participants in the first week cohort work in the tourism sector. This fraction reduces to 20-30% for the cohorts of weeks 2 and 3, and keeps decreasing further afterwards. The businesses participating from the tourism sector are broadly distributed, representing a comperatively larger fraction of businesses in the regions of St. Moritz, Davos, Pontresina and Arosa. Conversely, the other business sectors are more concentrated in Chur and to a less degree in Davos. In the first week cohort we observe a prevalence of individuals employed in companies located in Davos, St. Moritz, and Pontresina, as well as a noticeable fraction of individuals from peripheral regions with shared borders (e.g., Bregaglia). Instead, the cohorts of week 2 and week 3 are characterized by a higher representation of the region of Chur, which hosts offices and factories, in addition to the touristic region of St. Moritz which is also present in week 1 cohort (see *Supplementary materials, Cohort definition*).

**Figure 2:**
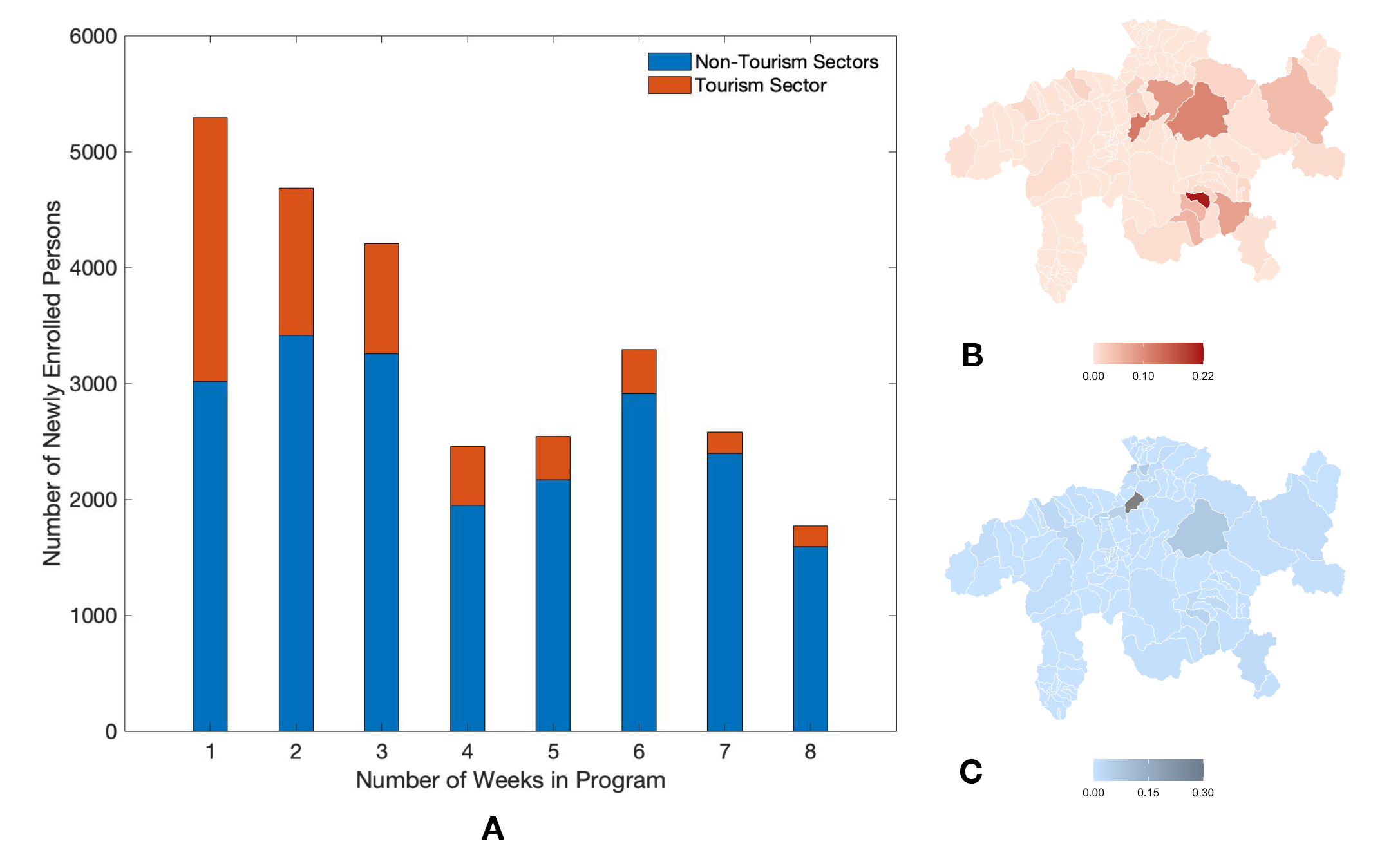
Enrollment and geographic distribution. Panel A: Number of newly enrolled employees versus the number of weeks since initiation of the program. Panels B and C: The geographic distribution of work-places of program participants in tourism sector and non-tourism sectors, respectively (see Fig 3 in *Supplementary materials* for more details).

The number of tourists peaked around December and February, and started declining towards the middle of March. This variation in the number of tourists could explain the increase of TPR among the newly enrolled individuals (control group without testing) during the month of February (peaking around the fourth week) and the decline observed afterwards. The tourists and the large proportion of foreign workforce employed in the tourism industry are a potential source of exogenous disease incursion and can noticeably contribute to the incidence of COVID-19 in the Canton Grisons. The fluctuation in the proportion of businesses from the tourism sector in the three cohorts is a potential source of confounding effect in our analysis.

In particular, the week 1 cohort bears the highest proportion of the tourism work force and shows the least reduction in TPR. This might be explained by two effects. First, the employees who were mostly in contact with visiting tourists were subject to a higher disease transmissibility, resulting from a larger number of contacts with untested individual (as tourists are not part of the program) and possibly coming from regions with higher prevalence. Second, many employees in this sector were crossing borders on a daily or weekly basis, hence their risk of exposure to infected individuals might differ from that of employees in non-tourism sectors, which mostly remained within the Canton during the time of our study. These hypotheses are supported by an analysis performed on a reduced data set from which a few touristic and near-border municipalities have been excluded. The reevaluated reduction in the incidence rate almost doubles from 18% to 34% for week 1 cohort, and increases from 48% to 51% for week 2 cohort, and from 43% to 61% for week 3 cohort (see Table 4 and *Supplementary material, Effect of tourism and cross border commuters* for the details).

**Table 2:** Ratio between the incidence rate of each repeatedly tested cohort with respect to the newly enrolled population, for unit cohort size of 3

| Starting week of cohort | Estimate | p-value | 95% CI |
| --- | --- | --- | --- |
| week 1 | 0.84 | 0.42 | (0.55,1.28) |
| week 2 | 0.48 | 0.01 | (0.26,0.85) |
| week 3 | 0.54 | 0.05 | (0.27,1) |

**Table 3:**
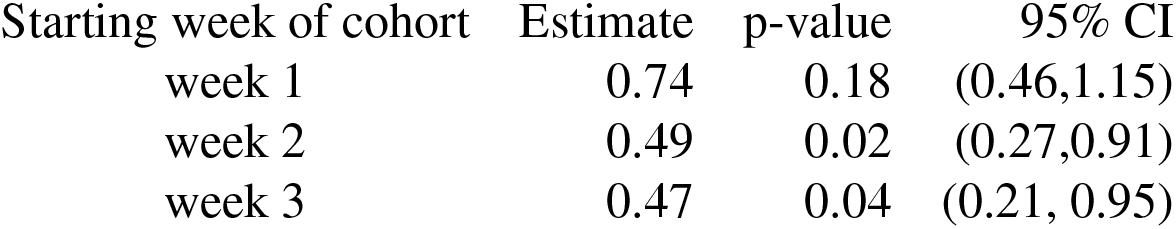
Ratio between the incidence rate of each repeatedly tested cohort with respect to the newly enrolled population, for unit cohort size of 3

**Table 4:**
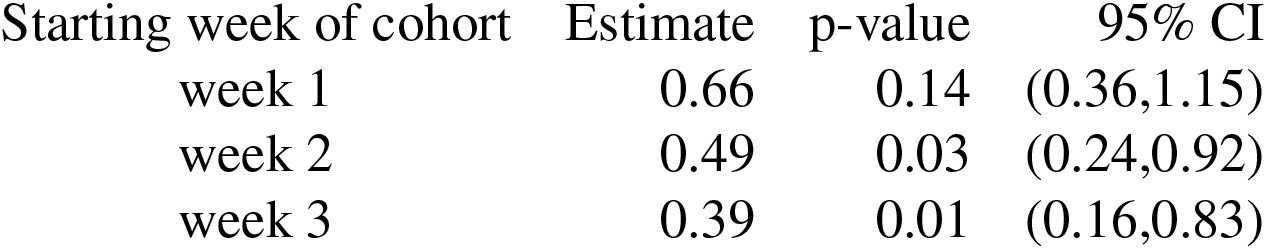
Ratio between the incidence rate of each repeatedly tested cohort with respect to the newly enrolled population, excluding touristic and peripheral regions

| Starting week of cohort | Estimate | p-value | 95% CI |
| --- | --- | --- | --- |
| week 1 | 0.66 | 0.14 | (0.36,1.15) |
| week 2 | 0.49 | 0.03 | (0.24,0.92) |
| week 3 | 0.39 | 0.01 | (0.16,0.83) |

## Discussion

The question whether repetitive mass testing can play a key role in response to COVID-19 pandemic, has important health, economical and social implications. The data collected from business testing program of the Canton Grisons provide the first empirical evidence that repetitive mass testing is indeed effective to prevent spread of the epidemic. Even though the repetitive testing campaign for COVID-19 mitigation on such a population scale and over such an extended period of time is unprecedented, our sample size remains relatively small: about 200 positive tests resulting from repetitive testing of around 27’000 individuals over eight weeks. This results in a considerable uncertainty in our analysis. Nevertheless the reduction in the incidence rate that we observed is statistically significant, especially among the groups of employees with lower exposure to exogenous disease incursions (e.g., individuals not employed in the tourism sector nor living in touristic/peripheral regions). This is encouraging as it shows that intensifying repetitive testing can be a viable complement to the existing non-pharmaceutical intervention strategies to reach epidemic control.

Several features contributed to the success of our testing program. The use of saliva based RT-PCR tests, with no nasal swab required, has certainly fostered a high participation rate and consistent repetitive testing: the program uptake among employees was 74% during February-March 2021, with an estimated drop-out by the second month lower than 10%. Furthermore, the high specificity of the employed RT-PCR tests assures that only a few false positive cases can be expected; this is important to keep testing and self-isolation effective also when the prevalence of the tested population is relatively low. Finally, the efficiency of the test analysis was improved by automated pooling of the samples in the laboratory, which allowed us to scale up the size of the tested population for a given testing capacity and to keep the test-to-notice time short.

To define the objectives and plan a successful repetitive testing concept, it is important to carefully consider the interaction within and between the tested and untested sub-populations, as their dynamic is far from being homogeneous. On the one hand, the repeatedly tested individuals actively contribute to curb the virus spread: as the tested cohort itself benefits from a reduced incidence, their untested contacts are also exposed to a lower risk of infection. On the other hand, interactions of tested cohort with individuals outside of the program expose the former to the transmission risk originated from the latter, reducing the effectiveness of testing.

Our analysis supports that a higher number of active participants per company site tended to correlate with smaller incidence rates, hence confirming the requirement of achieving a sufficiently large uptake among employees (*Supplementary materials, Overview of statistical methods*). Also, a higher number of tourists or cross-border employees from regions not covered by the testing program represented a potential source of disease incursion and were associated with a lower reduction of the incidence rate. In order to more efficiently control the epidemic, therefore it is important to adapt the testing program according to the varying level of exposure to the exogenous sources of infection. This can be achieved, for instance, by a larger uptake and a more frequent testing among individuals with an extended network of contacts outside of the program.

If carefully designed, repetitive mass testing programs can complement other public health measures, limiting socio-economical tolls resulting from extreme interventions. In the current COVID-19 pandemic, as the share of vaccinated people is growing fast across several countries, including Switzerland, a certain amount of repetitive testing can still help complement the relaxation of measures until a sufficiently large fraction of the population is vaccinated. Resorting to repetitive mass testing programs remains certainly valuable, for instance, in societal branches for which vaccination isn’t possible yet or in which outbreaks due to exposure to new mutants is highly probable (e.g. at schools or in the tourism sector). This requires a coordinated engagement of policy makers, communities and the public to adopt carefully designed testing programs that could be sustained over an extended period of time.

## Data Availability

The source data set is available on https://www.gr.ch/DE/institutionen/verwaltung/djsg/ga/coronavirus/info/medien/Seiten/Medien.aspx. The analysis was implemented with R. The corresponding codes are available upon request from the corresponding author.

## Supplementary materials

Methods

Figs. 3 to 4

Tables 2 to 4

## Methods

### Study design and data acquisition

After the company signs up for the testing program, the employees register to the IT system and the validity of their data entry is confirmed. Saliva samples are collected by rinsing the oral cavity with 2 ml reagent grade water for one minute. Program participants are instructed not to eat, drink, smoke or brush teeth 1 hour before sample collection. The fluid is then collected by means of a plastic funnel into a dedicated CE-marked sterile sample tube containing crystalline guanidine thiocyanate in order to inactivate the sample and stabilize viral RNA. We determined stability of RNA-levels at room temperature for 5 days. Testing vessels are delivered by the Swiss post. For the collection of the samples, the employer delivers the samples to predefined collecting points from where the Swiss post together with the regional train, transport the samples to the lab.

Pools of 5 samples received at the same day in the laboratory are mixed in a fully automated platform preventing contamination in the pipetting procedure. For each single program participant an additional separate sample is produced and stored in a sample archive. RNA of the pool sample is purified by means of an automated extraction platform. The SARS-CoV-2 specific RNA is then amplified by means of a commercial test kit run on a QuantStudio 5 instrument. This assay tests for presence of 3 different genes of the SARS-CoV-2 virus (N-gene, ORF1abgene, S-gene). Negative and positive control materials are tested with each run. When a pooled sample tested positive or ambiguous, the stored samples of the pool participants are analyzed at an individual level, in order to identify individuals positive for SARS-CoV-2. The results are communicated within 24 hours upon submission of the samples.

Positively identified individuals are asked to self-isolate for ten days. The quarantine of the work contacts of the positive cases is waved and replaced by daily testing for maximum of ten days; the work contacts of the positive case are asked to self-isolate only in case their test results turns out positive. Test results from the laboratory are entered into the IT platform by hand and the results are expressed in MS Excel spreadsheets. The pseudonymized data set includes unique identifiers for the company site and the tested employee, besides the testing date, the test result, the municipality of the company and the business sector.

### Data pre-processing

From 146’823 entries related to company testing in February and March 2021, 25’449 (17%) entries did not have a test result. This was typically the case when an employee gets a testing voucher but does not take the test. We remove the records without test results, and analyze only the curated data set.

In the case of multiple tests per seven-day time window, only one test per person is counted. The result of the test is considered positive if any of the tests taken in the seven-day time window gives a positive result.

The contacts of positive cases comprise a high-prevalence sub-population, thus the chance that they are identified as positive is higher than for a randomly chosen person from the same cohort (as contacts are tested daily). On the other hand, excluding the contact testing records from the analysis would make the number of positive cases in repeatedly tested cohorts biased towards lower values (as the contacts of positive cases would not be counted). Therefore we make a conservative choice and do not differentiate between the test results of the contact testing program from the rest. We expect that an ideal random testing program would reach a stronger effect compared to the results of our analysis.

In total, 266 tests rendered positive results, yet 51 are repeated positive tests (19% of all positive tests). This is due to three main reasons:

1. The delay in the test-to-notice process for daily contact testing can lead to a scenario where a person is tested on the following day while still uninformed about positive result of the past day. As a result, two positive tests of the same person in two consecutive days would be recorded in the data set.
2. After recovery, a person might still show up positive in the PCR test.
3. In a few cases, the positive tested employee does not adhere to the self-isolation rule and repeats the test.

In our analysis, which covers a period of two months, we count a positive test result only once per person.

### Cohort definition

We define the following sub-populations of test results.

1. Newly enrolled population: A test is counted in the newly enrolled statistics if the person hasn’t been previously tested in our testing campaign, and if the company site has not been in the program for more than two weeks prior to the test date. The latter condition is introduced to exclude (to some extent) the effect of community protection due to testing, which might occur among those employees of a company that have started to participate later.
2. Week 1-3 cohorts: These cohorts are composed of employees that enter the program during the same week (numbered sequentially from the start of the program) and named week 1, week 2, and week 3 cohorts. During its first seven days each cohort coincides with the newly enrolled population. We only count an individual in the cohort if at least 5 persons from the same company site participate consistently (i.e., they are tested every week). In other words, we only keep those individuals in the cohort, whose companies sites remain active in the program. Notice that the results are robust with respect to the adopted minimum number of active participants per company site, henceforth called “unit cohort size” (illustrated in Tables 2 and 3).

Figure 3 shows the geographical distribution of the work places of newly enrolled employees. The share of touristic and near-border regions is noticeably higher in the week 1 cohort than in the cohorts of the two subsequent weeks.

The program participation rate during February-March 2021 has been 74%; 9.9% of employees who tested at least once during February, submitted no test during March, suggesting an estimated drop-out rate of 10% per month.

### Overview of statistical methods

The daily data are too scarce and largely affected by the weekly periodicity, particularly visible in the consistently lower number of tests during the weekend. Therefore, we apply a moving average with a seven-day time window (forward in time). The following two analyses are then performed.

1. Test Positivity Rate (TPR) : To compute the seven-day average TPR at a given date in a certain sub-population, the total number of discovered positive cases is divided by the number of tests in the seven-day window. Only one test per person is counted per considered period. If *τ* is the time (day), let *Ī*(*τ*) and 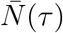 be the total number of new positive cases detected and the number of tested persons during the period [*τ, τ* + 6], respectively. Therefore the seven-day-average TPR is given by 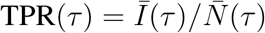. The histograms in Fig. 4 (A), (B) and (C) show the resulting TPR for newly enrolled employees side by side with the week 1,2 and 3 cohorts, respectively. For smoothing and uncertainty quantification of time series data, we follow an approach similar to the one discussed in (*15, 16*). The seven-day average TPR is smoothed using a first order local polynomial regression (Loess) algorithm with bandwidth 0.65 (*17*). To quantify the uncertainty band, we apply bootstrapping prior to the computation of the seven-day average TPR and its corresponding Loess fit. A resampled record is obtained by sampling entries from the original data set under uniform probability and replacing them in the data set (*18, 19*). Thus the obtained resampled data set has a length similar to the original one. We generated 500 resampled data sets, and applied normal distribution for quantifying uncertainty of the estimated statistics. To ensure that the starting points of week 1,2 and 3 cohorts and their uncertainties lie on the ones obtained from the newly enrolled population after smoothing, the Loess weights are locally adjusted and the neighbourhood of the intersection points are smoothed again by moving averaging. The Loess fitted TPR and its 95% CI are shown in Fig. 1 (B), (C) and (D) for week 1,2 and 3 cohorts, respectively, besides the newly enrolled population.
2. Incidence Rate Ratio: In order to extract information with a lower uncertainty than the TPR time series, we complement the analysis by calculating the incidence rate as an integral measure over the entire considered period. Accordingly, the following analysis would not rely on smoothing the seven-day average time series, and thus the results are independent of the Loess fit. We compute the incidence rate for each sub-population as proportion of the total number of new positive cases to the person-time, i.e. each person is counted for the number of weeks being tested in the sub-population. Therefore, the adopted incidence rate is an approximation of the area below the TPR curve (normalized by time). In order to avoid complications in our incidence rate estimations, we assume unity sensitivity and specificity of the tests. Furthermore, the time lag between infection and detection via the test is neglected. By binning the data of each cohort into weeks, we estimate the incidence rate of cohort 1 from week 2 till 8, cohort 2 from week 3 till 8, and cohort 3 from week 4 till 8. The incidence rates are compared with the incidence rate of the newly enrolled population for the same time period.

**Figure 3:**
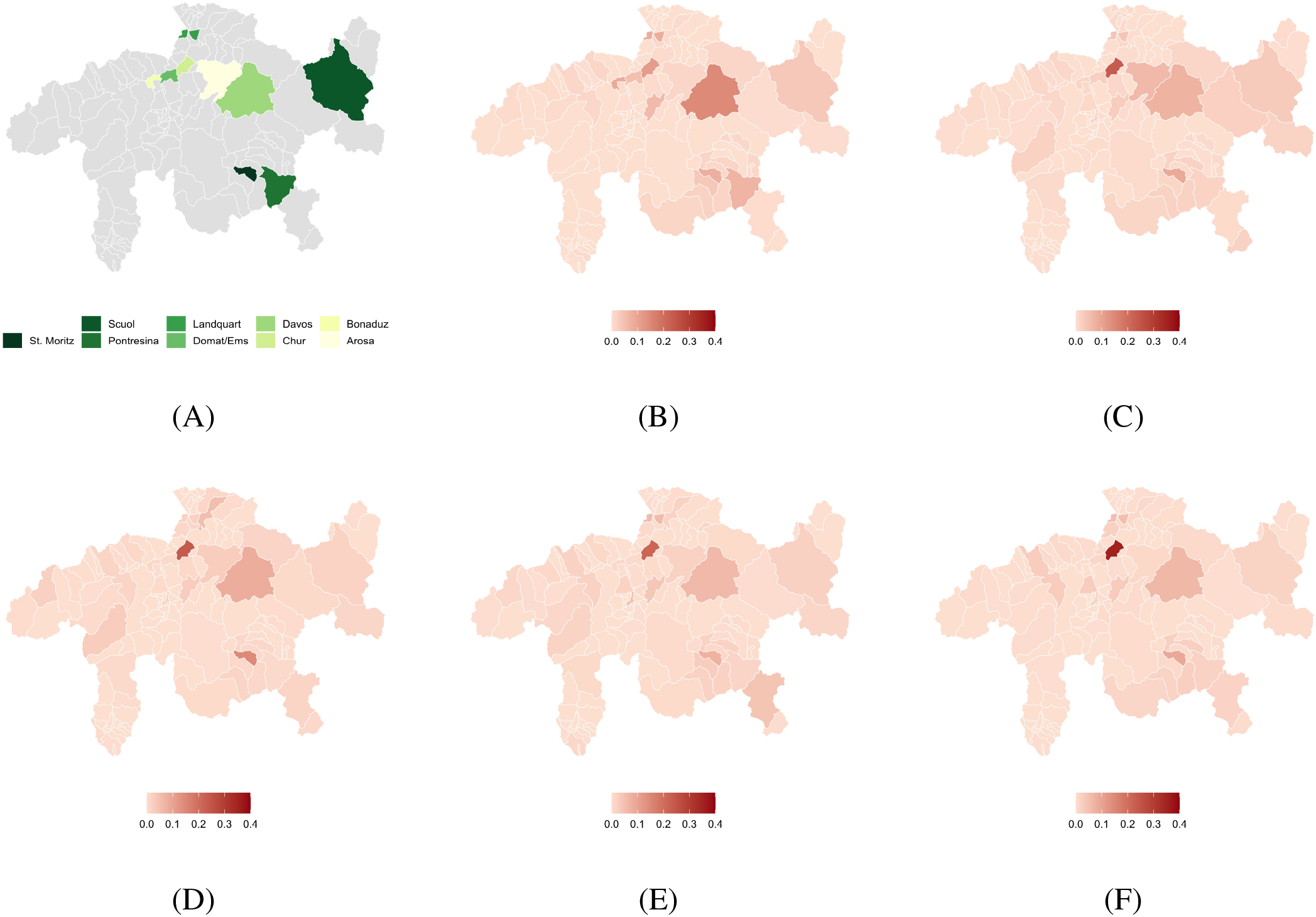
Geographical distribution of the participating businesses. Enrollments from week 1 to week 5 correspond to panels (B) to (F), respectively. The distribution of the work place location of newly enrolled employees is color-coded for each week. Panel (A) provides names of few municipalities in the Canton Grisons.

**Figure 4:**
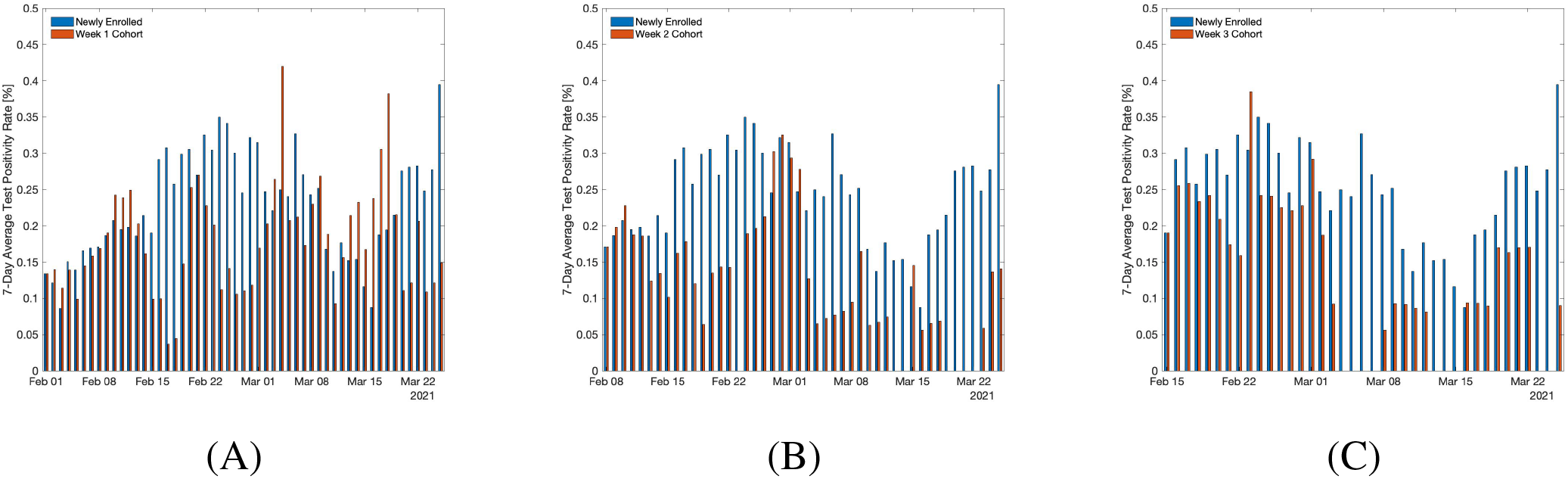
7-day average TPR (%). Panels (A), (B) and (C) show the TPR for newly enrolled employees side by side with the week 1,2 and 3 cohorts, respectively.

Consider 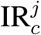 to be the incidence rate for the repeatedly tested cohort that joins the program at week *j ∈ {*1, 2, 3*}*. Let *N*^*j*^(*i*) and *I*^*j*^(*i*) be the number of tested people and number of new positive cases, respectively, at week *i* in the cohort *j*. Therefore the incidence rate for a cohort that starts at week *j* is

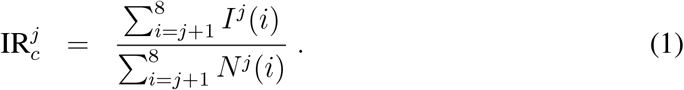

Accordingly, for the newly enrolled incidence rate IR_0_ we have

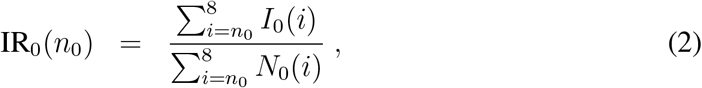

where *N*_0_(*i*) and *I*_0_(*i*) are the number of tested people and discovered positive cases, respectively, at week *i* among the newly enrolled population. In order to compare the incidence rate of each cohort with the newly enrolled population for a similar time interval, we set *n*_0_ to 2, 3 and 4, for comparing IR_0_ to 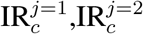 and 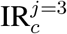, respectively. The corresponding reduction in the incidence rate for a cohort starting at week *j* then follows

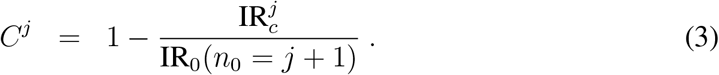

The reduction in the incidence rate is computed from the incidence rate ratio estimated by the package Epitools (*20*). The confidence intervals together with p-values are computed using mid-p exact method (*20, 21*). The results are provided in Table 1. Furthermore, we repeat the evaluation of the incidence rate ratio by changing the minimum number of consistent participants per company site. The results are given in Table 2 for unit cohort size of 3, and in Table 3 for unit cohort size of 7. While the estimates do not vary much with respect to the unit cohort size, we observe that larger unit cohort sizes tend to correlate with smaller incidence rate ratios, and thus stronger reduction in the incidence rate. For example in the week 1 cohort, we find that the reduction increases from 16% to 26%, once the unit cohort size grows from 3 to 7. This is in line with the expectation that more effective reduction in the incidence rate would be achieved in companies, where larger pools of employees participate in the program.

### Effect of tourism and cross-border Commuters

To better understand the disparity in TPR and incidence rate reduction of different cohorts, we analyze the effects of tourism-sector employees and cross-border commuters. While it is possible to exclude from the data set the employees defined as working in the tourism sector and reevaluate the effect of repetitive testing in the filtered data set, it may not provide a correct evaluation of the effects of visitors and commuters. Indeed, not all business branches that deal with tourist clients are categorized as tourism sector (e.g., employees that provide various services in train stations interact often with tourists but are not counted as being employed in the tourism sector); moreover, our cohorts are not isolated and interact with large portions of the population outside of the program. Also, due to the limited amount of data (resulting in wide uncertainty bounds, Fig. 1), removing a large portion of the data can be misleading. It may be more appropriate to take into account tourist-related infections by performing a regional analysis of the data rather than separating the different business sectors. Therefore, we decided to exclude a few touristic and near-border municipalities from the data set.

In the week 1 cohort we identify three regions with the highest number of employees working in the tourism sector: Davos, Pontresina and Vaz/Obervaz. We remove from the data set these municipalities as well as two peripheral regions Landquart and Bregaglia, which are more represented in the week 1 cohort than in the other two. In the filtered data set we observe that the share of tourism sector is shrank from 43% to 34% in week 1 cohort, from 27% to 26% in week 2 cohort, and from 23% to 22% in week 3 cohort. The incidence rate ratio of each cohort with respect to the newly enrolled one is re-evaluated and reported in Table 4. The incidence rate reduction almost doubles from 18% to 34% for week 1 cohort, and increases from 48% to 51% for week 2 cohort, and from 43% to 61% for week 3 cohort. This supports the hypothesis that the relatively small reduction of TPR of the week 1 cohort in the original data set can be explained by an over representation of regions with higher rates of tourists and cross-border commuters.

## Data and Code Availability

The source data set is available on https://www.gr.ch/DE/institutionen/verwaltung/djsg/ga/coronavirus/info/medien/Seiten/Medien.aspx. The analysis was implemented with R. The corresponding codes are available upon request from the corresponding author.

## Notes

### Competing Interest Statement

Lorenz Risch and Martin Risch are key shareholders in Risch laboratories, which provided the laboratory testing.

### Funding Statement

No external funding was received

### Author Declarations

The study was reviewed by the Cantonal Research Ethics Commission of Zurich Switzerland (KEK-ZH), and obtaining ethical approval was waived.

